# Exploring the local field potential signal from the subthalamic nucleus for phase-targeted auditory stimulation in Parkinson’s disease

**DOI:** 10.1101/2023.10.31.23297547

**Authors:** Elena Krugliakova, Artyom Karpovich, Lennart Stieglitz, Stephanie Huwiler, Caroline Lustenberger, Lukas Imbach, Bartosz Bujan, Piotr Jedrysiak, Maria Jacomet, Christian R. Baumann, Sara Fattinger

## Abstract

**Background:** Enhancing slow waves, the electrophysiological (EEG) manifestation of non-rapid eye movement (NREM) sleep, might offer therapeutic benefits for patients with Parkinson’s disease (PD) by improving sleep quality and slowing disease progression. Phase-targeted auditory stimulation (PTAS) is an approach to enhance slow waves, which are detected in the surface EEG signal in real-time.

**Objective:** We aimed to test whether the local-field potential of the subthalamic nucleus (STN-LFP) can be used to detect frontal slow waves and evaluate the PTAS-related electrophysiologic changes.

**Methods:** We enrolled patients diagnosed with PD and undergoing Percept™ PC neurostimulator (Medtronic) implantation for the deep brain stimulation of STN (STN-DBS) in a two-step surgery. Patients participated in 3 full-night recordings, including one between-surgeries recording and two recordings during rehabilitation, one with DBS+ (on) and one with DBS-(off). Surface EEG and STN-LFP signals from Percept PC were recorded simultaneously, and PTAS was applied during sleep in all three recording sessions.

**Results:** Our results show that during NREM sleep, slow waves of the cortex and STN are time-locked. We observed the effects of PTAS in STN-LFP in all types of analyses, including spectral, coherence, and event-related potentials.

**Conclusion:** Our findings suggest that PTAS could be implemented using STN-LFP signal for slow wave detection. Additionally, we propose options for better STN-LFP signal preprocessing, including different referencing and filtering that could make the detection of the cortical slow waves in STN-LFP more reliable.

## 1. Introduction

In today’s ageing world population, neurodegenerative diseases pose significant medical challenges as they become increasingly prevalent. Still, curative or disease-modifying treatments are not available, despite some promising new strategies in the field of Alzheimer’s disease. Parkinson’s disease (PD), one of the fastest-growing neurodegenerative disorders worldwide (Feigin et al., 2017), currently affects 1-2% of individuals aged 60 and above (Pringsheim et al., 2014). Besides the cardinal motor symptoms of PD (i.e., akinesia, tremor, rigidity, postural instability), sleep-wake disturbances, including poor nighttime sleep and excessive daytime sleepiness, affect up to 80-90% of patients, significantly worsening their quality of life (De Cock et al., 2008). Growing preclinical and clinical evidence indicates that improving non-rapid eye movement (NREM) sleep may slow down the progression of neurodegenerative diseases such as Alzheimer’s disease and PD (Gerstner et al., 2012; Roh et al., 2012; Noble and Spires-Jones, 2019; Morawska et al., 2021). In particular, the unique neuronal firing pattern of the thalamocortical network during NREM sleep, registered by the surface electroencephalogram (EEG) as slow waves, has been linked to synaptic and neuronal homeostasis (Vyazovskiy and Harris, 2013; Tononi and Cirelli, 2014), as well as the removal of waste products through increased glymphatic flow (Xie et al., 2013; Hablitz et al., 2019). Moreover, in a retrospective study of 129 PD patients, our group found an inverse correlation between the cumulative sum of slow waves during the examined night and subsequent motor symptoms progression (Schreiner et al., 2019). Thus, boosting slow waves during NREM sleep may not only exert immediate therapeutic effects on vigilance and sleep quality in PD (Büchele et al., 2018) but may also have disease-modification effects on neurodegeneration. Despite the clear benefit of improvement of NREM sleep in PD, pharmacological enhancement of slow waves is burdened with side effects and bears the risk of developing tolerance and dependence. There is, however, a promising non-pharmacological approach to increase slow waves: phase-targeted auditory stimulation (PTAS). In a seminal study of Ngo et al., (2013) it was shown for the first time that slow waves can be specifically enhanced by presenting non-arousing auditory stimuli phase-locked to the ongoing slow waves. Furthermore, such slow waves enhancement led to improved sleep-dependent behavioral performance (Ngo et al., 2013). Subsequent studies have offered additional evidence supporting PTAS as a novel method to modify slow waves (Fattinger et al., 2017; Krugliakova et al., 2022), potentially benefiting patient populations experiencing diminished sleep-related recovery (Papalambros et al., 2019; Prehn-Kristensen et al., 2020), including older adults (Lustenberger et al., 2022).

Deep brain stimulation of the subthalamic nucleus (STN-DBS, Fig. 1), is a gold-standard treatment in PD (Sharma et al., 2016). STN-DBS consists of a subcutaneously placed neurostimulator connected to the electrodes implanted bilaterally into the STN, continuously applying electrical impulses (Benabid et al., 1991, 1994; Siegfried and Lippitz, 1994). Such DBS treatment improves motor symptoms, reduces PD-related pain, and benefits neuropsychiatric fluctuations and impulse control (Lhommée et al., 2012, 2018; Sürücü et al., 2013; Baumann-Vogel et al., 2015). Importantly, it also improves subjective sleep quality and reduces daytime sleepiness (Baumann-Vogel et al., 2017; Sousouri et al., 2021). Only recently Medtronic has introduced a new neurostimulator Percept™ PC, which allows not only the application of electric impulses but also to simultaneously measure local field potential (LFP) of STN in real-time. Thus, this new technological solution within a standard patient treatment, allowing capturing STN-LFP, offers an opportunity for integrating of PTAS into DBS for PD patients without the need for an additional external EEG device. In line with our aspirations, a recent study in epilepsy patients has shown the feasibility of phase-targeted stimulation approach to influence sleep-specific EEG rhythms based exclusively on the intracranial recording (Geva-Sagiv et al., 2023).

**Figure 1.**
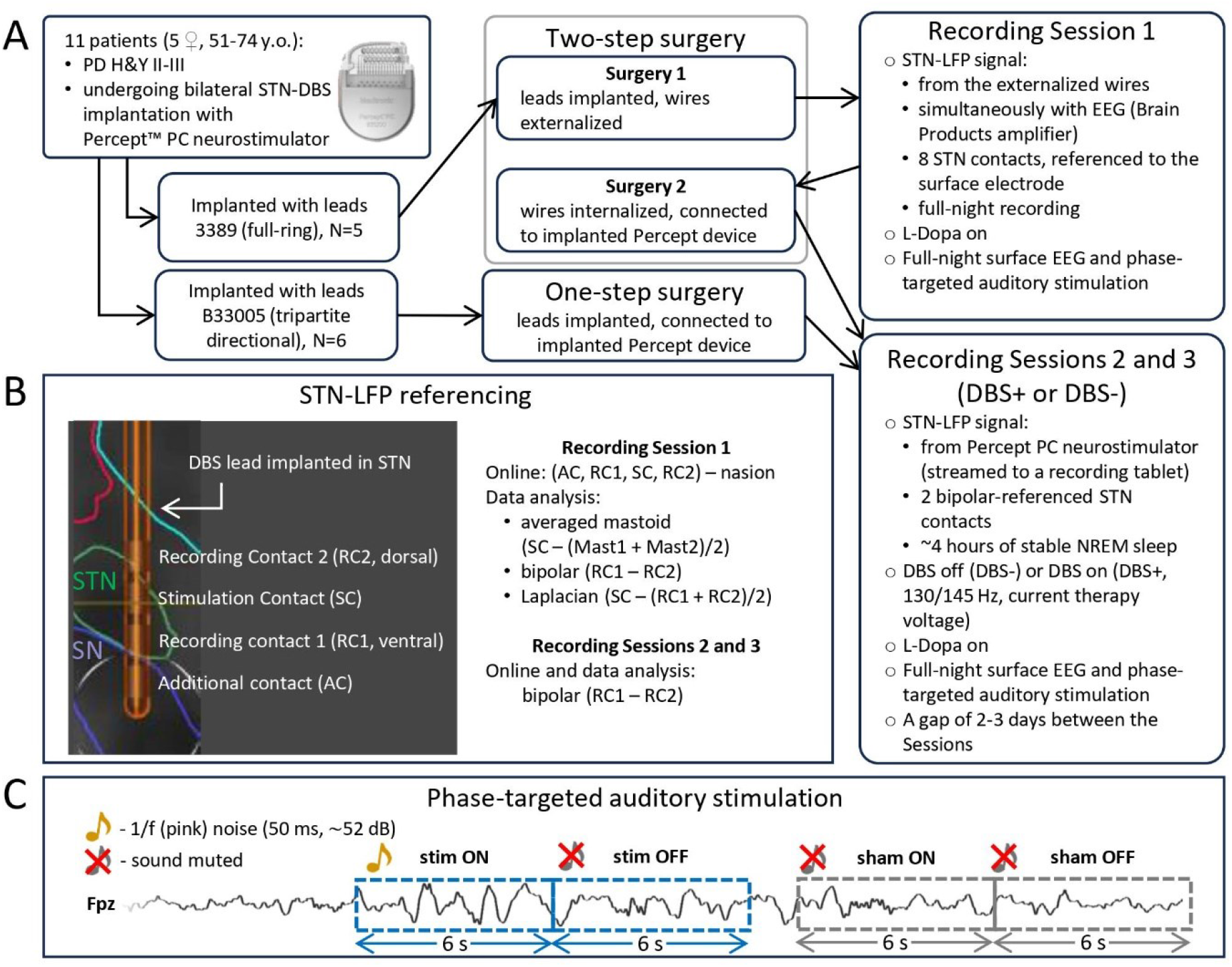
Experimental design and data recording. **(A) Study flowchart**. Whether participant will take part in Recording Sessions 1 (between two surgeries) depended on the type of leads implanted. Recording Session 2 and 3 (DBS+/DBS-) took place for all participants during the rehabilitation period. The order of DBS+ and DBS-sessions was randomized and counterbalanced. **(B) Reference montage used for STN-LFP recording and data analysis**. Each DBS lead has 4 contacts. Based on the postoperative reconstruction of the electrode placement, contact in the dorsolateral STN was selected as Recording Contact. Two neighboring contacts were used for the bipolar recording (Recording Contacts). **(C) Phase-targeted auditory stimulation protocol**. During all Recording Sessions, full-night surface EEG was recorded, and up-phase-targeted auditory stimulation was applied throughout the night. The signal of electrode Fpz was used for the automatic detection of NREM sleep episodes and suitable slow waves within that period. During auditory stimulation, pink noise pulses were delivered via in-ear earphones (Etymotic Research Inc., ER 3C) in 6-s blocks (stim ON-windows), followed by a 6-s pause (stim OFF-windows); sham ON and sham OFF windows were identical to stim condition, but the sound was muted. Conditions order was pseudorandomized. Abbreviations: PD – Parkinson’s Disease, H&Y II-III – Hoehn and Yahr scale (stages 2 and 3), STN-DBS – deep brain stimulation of the subthalamic nucleus, LFP – local field potential, RC – recording contact, SC – stimulation contact, AC – additional contact, STN – subthalamic nucleus, SN – substantia nigra.

The overall goal of this study is to test the compatibility and feasibility of the integration of the PTAS into STN-DBS therapy by assessing whether STN signal can be used for detection of surface EEG slow waves. We demonstrate that the usual (temporal and spectral) profile of delta-range activity (slow waves frequency band) can be detected during NREM sleep within the STN using implanted DBS leads. We observed that STN-LFP activity in delta range is coherent with cortical slow waves as registered by surface EEG, but that the possibility to detect slow waves in STN is strongly affected by the reference method. Furthermore, we show that a response to PTAS, like delta power increase, can be captured in STN-LFP.

## 2. Methods

### 2.1. Patients and surgical procedures

Electrophysiological data was collected in 11 patients (5 females, 51-75 years old) diagnosed with mild/moderate PD (Hoehn-Yahr stages ll-lll) undergoing bilateral STN-DBS implantation at the Department of Neurosurgery, University Hospital Zurich, Switzerland. For more information about patient’s characteristics, the screening night and other preoperative inclusion criteria, see Supplementary Methods. The study protocol was approved by the local ethics committee (Kantonale Ethikkommission Zürich, KEK-ZH, project number 2021-01327). All patients provided written informed consent.

Patients were implanted bilaterally with DBS leads in STN. Five patients were implanted with Medtronic (Medtronic Neurological Division, Minneapolis, MN, USA) lead model 3389 (addressed after as full-ring leads), and 6 with model B33005 (tripartite directional leads). In the case of full-ring leads, the internalization and connection of the lead wires to the stimulation device (Percept™ PC, Medtronic Neurological Division, Minneapolis, MN, USA) was performed 3–5 days after the first surgery (two-step surgery) (Fig. 1, A). Thus, direct recording from the externalized DBS leads (i.e., RS1) was possible only in 5 patients implanted with full-ring leads (for more details, see Supplementary Methods, Surgical procedure section).

### 2.2. Experimental protocol

#### 2.2.1. Recording sessions and STN-LFP recordings

After the surgery, sleep was recorded simultaneously with EEG and STN-LFP in all 3 Recording Sessions and PTAS was performed based on slow waves detected in the frontal surface electrode (Fig. 1, A). Patients took their regular medication during all 3 Recording Sessions (i.e., L-Dopa on, see Supp. Table PC). Recording Session 1 (RS1, with full-ring leads only) was performed one or two nights after the DBS leads implantation. During RS1, data were collected in DBS-(OFF) state and prior to the implantation of the Percept device (second surgery). The STN-LFP was recorded from both hemispheres using externalized wires simultaneously with EEG, connected through a touch-proof adapter to the BrainAmp DC amplifier. Along with other surface EEG channels, all 8 STN contacts were recorded referenced to the frontal surface electrode.

During the rehabilitation period following the completion of the surgery, Recording Sessions 2 (RS2) and 3 (RS3) took place (order randomized and counterbalanced), with a gap of 2-3 days. Simultaneously with the surface EEG, STN-LFP recording was performed with implanted Percept PC neurostimulator (250 Hz sampling rate, built-in filters: low-pass at 100 Hz and high-pass at 1 Hz) and transmitted wirelessly to the dedicated tablet (BrainSense™, Streaming mode). The STN-LFP recording was possible only for the first part of the night (approximately 4 hours of NREM sleep) due to the constraints of the recording system. Throughout one of the nights, DBS was turned off (DBS-), while during another DBS was on (DBS+), the order of these nights was counterbalanced across participants. During DBS+ night, monopolar STN-DBS was delivered at 130/145 Hz with a current therapy voltage through the electrode contact positioned in the optimal functional target (see Supplementary Table AT). Data were always recorded with a bipolar referencing montage from 2 contacts, neighboring to the stimulation contact: the signal of more dorsal contact was subtracted from the more ventral contact (see Supplementary Table CL, and Fig. 1, B).

#### 2.2.2. Surface EEG recording and phase-targeted auditory stimulation

Full-night sleep EEG, electrooculogram (EOG), and electromyogram (EMG) was recorded using a standard 10-20 system (Grass Gold Cup Electrodes, Natus Neurology Inc, Warwick, USA). For the continuous recording of the EEG and LFP (only during RS1), a BrainAmp DC amplifier (Brain Products GmbH, Gilching, Germany) was used. EEG was referenced to nasion electrode and recorded with a sampling frequency of 500 Hz. All electrode impedances were below 20 kΩ at the start of the recording.

To reliably estimate the PTAS effect, we used a paradigm with two conditions (stim and sham), each comprising two subsequent 6-sec windows (ON and OFF, Fig. 1, C). In the stim ON condition, PTAS was applied phase-locked to frontal slow waves (targeting zero-to-up transition of slow wave in Fpz channel). OFF windows were included in the design to provide a washout time for the PTAS effect (Krugliakova et al., 2022). For the sham condition, PTAS was muted but trigger positions were saved. PTAS algorithm was implemented in OpenViBE (described in Huwiler et al., 2022; see Supplementary Methods, section Phase-Targeted Auditory Stimulation).

### 2.1. Data preprocessing and analysis

Nine out of the 11 patients had sleep efficiency more than 70% either during RS1 or both RS2 and RS3 and were included in the analysis. A summary of the data recorded in each of these 9 patients and clarification which patient was included in which analysis is presented in Supplemetary Table AT. EEG and LFP preprocessing and analysis was performed in MATLAB R2019a (The MathWorks, Inc., Natick, MA), using FieldTrip toolbox (Oostenveld et al., 2011) and custom-written scripts.

As embedded synchronization input/output signals are not presently available for the Percept PC, the RS2 and RS3 data was manually synchronized with the EEG prior to the data processing (see the Supplemetary Methods). For one of the patients, cardiac artifacts were deleted from the LFP data using brMEGA algorithm (Chiu et al., 2022, see Supp. Table AT).

All data (except for the Percept LFP streams) were first high-pass filtered at 0.5 Hz (one-pass, windowed-sinc zero-phase FIR filters, Hamming window; order 1,650, cutoff −6 dB at 0.5 Hz). Subsequently, all data were low-pass filtered at 45 Hz (same as high-pass, order 148, cutoff −6 dB at 45 Hz), downsampled to 250 Hz. EEG was re-referenced to the average mastoid. One of the goals of our study was to investigate how referencing affects the similarity of LFP signal to the surface EEG, and for RS1 LFP recording, we probed three referencing montages: (1) *averaged mastoid referencing* (captures large-amplitude events), (2) *bipolar referencing* (montage used in Percept device), (3) *Laplacian referencing* (highlights the local STN activity) (see Fig.1, B). To increase the number of observations for the statistical analysis, we included each STN as a separate observation. This resulted in N=10 (5 patients * 2 STNs) for each RS. To minimize the influence of the individual signal amplitude, before dividing STNs, all data were z-score transformed within each patient.

EEG and LFP data analysis was performed for the ON-OFF pairs, that belonged to manually scored N2-N3 sleep stages (see Supplemetary Methods, section Sleep Scoring) and did not have artifacts identified manually using *ft_rejectvisual* (FieldTrip toolbox). The offline wave detection was performed for stim OFF, sham ON and sham OFF windows (all window types where the sound was muted) of RS1. Slow waves positive peaks were detected in Fpz data filtered between 0.5 and 2 Hz, the duration of half-wave (positive-to negative-zero-crossing) should have been 0.25-1 s. To improve the signal-to-noise ratio, and because larger slow waves tend to be global events, only the top 30% largest waves of each participant were included in the analysis (mean=830, min=133, max=1884; of note, similar results were found when all slow waves were included in this analysis). Time-resolved power (offline wave detection and ERP/ERSP analysis) was calculated using wavelets with a Hanning taper and an adaptive time window for each frequency (*ft_freqanalysis* function, *mtmconvol* method in FieldTrip toolbox). Power was estimated in the band between 4 and 25 Hz (5 full cycles per window, ΔT =5/f) in steps 0.02 s and 0.25 Hz frequency bins. Power for the spectrogram was assessed in 6-s ON-OFF windows (*mtmfft* method for *ft_freqanalysis* function in FieldTrip toolbox, Hanning taper, 0.5 to 18 Hz, 0.25 Hz frequency bins). To estimate synchronicity between the surface and STN-LFP, we performed coherence analysis for Fpz and STN-LFP using the same ON-OFF pairs as the power analysis (*ft_connectivityanalysis* in FieldTrip toolbox). Statistical analyses were carried out in Matlab R2019a and R studio. The statistical significance level (alpha) was set to 0.05 for all tests. To correct for multiple comparisons, we used a non-parametric clustering procedure (Maris and Oostenveld, 2007; Maris, 2012). For more details on the cluster correction and ERP/ERSP analysis, see Supplementary Methods.

## 3. Results

We present the findings from the three Recording Sessions in which we examined the potential for detecting slow waves registered in surface frontal EEG in the STN-LFP recording. Additionally, we evaluated the effects of PTAS and DBS status on the dynamics of surface EEG and STN-LFP signal.

### 3.1. Offline waves detection

In the analysis of RS1, we used the opportunity to reference the STN-LFP in three different ways, and investigated the effect of mastoid, bipolar, and Laplacian reference montages on detectability of cortical slow waves in STN-LFP.

To ensure that mastoid electrodes do not capture large-amplitude slow waves, and thus could influence the amplitude of slow waves observed in STN-LFP data when referenced to mastoid in RS1, we conducted an exploratory analysis in one of the patients. In short, no time-locked slow waves were observed for the mastoid electrode, and we further assumed that mastoid is a valid external reference location (for more information, see Supplementary Fig. SWMC).

To explore the synchronicity between the surface and STN-LFP slow waves, we initially focused on the windows of RS1 where no PTAS was applied (Fig. 2). Offline detection of the positive peaks of slow waves was performed in channel Fpz, referenced to the joined mastoid, while the STN-LFP signal (in three different reference montages) was time-locked to these peaks. The STN-LFP signal referenced to mastoids have shown higher phase precision (1/std=0.86±0.1, mean±std) as compared to both bipolar (1/std=0.78±0.04) and Laplacian (1/std=0.78±0.06) referenced data (both t(9)>2.34, p=0.04). While the mastoid-referenced STN-LFP has shown a phase reverse in relation to the Fpz-detected slow waves (except for one patient), the scattering of the resulting phase was higher for the other two reference montages. It is worth noting that low sigma power bursts, associated with transition to down phase of the surface slow waves, were present for mastoid- and bipolar-referenced STN-LFP, but not in the Laplacian montage.

**Figure 2.**
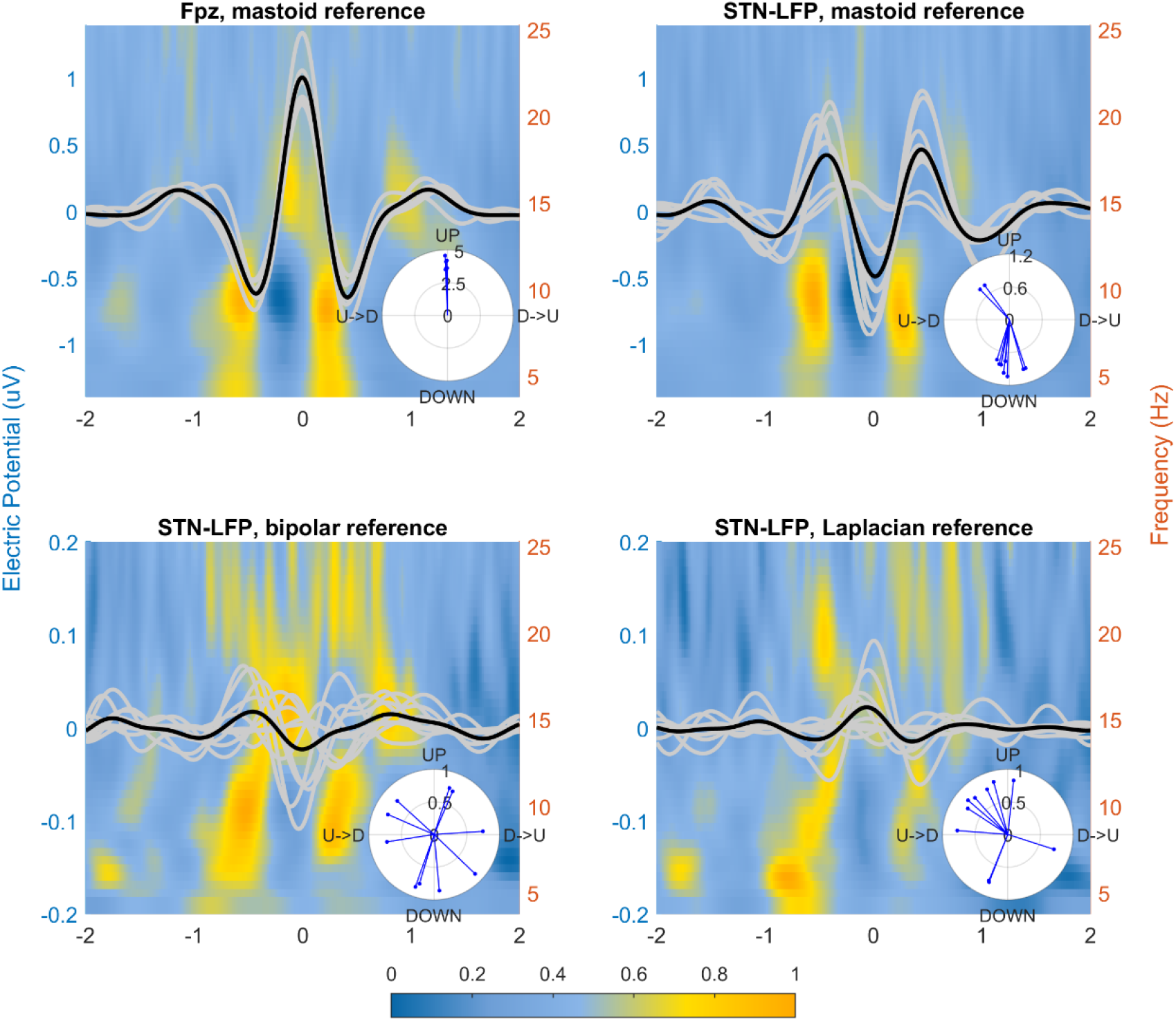
Average of offline-detected slow waves in the Fpz channel and STN-LFP signal time-locked to the peak of these cortical waves (Recoding Session 1, different reference montages). STN-LFP data is presented for three reference montages: average mastoid, bipolar, and Laplacian. The thick black line in each plot corresponds to the mean of all datapoints, gray lines – to separate datapoints (N=5 for Fpz and N=10 for STN-LFP). Inset polar plot shows the average phase at time point 0 (peak of slow wave in Fpz) for each datapoint, length of each vector indicates the phase precision (1/std); U->D indicates up-to-down transition, D->U – vice versa. The colourbar shows the normalized power (a.u.) of the background time-frequency plot.

### 3.1. Sleep macrostructure and number of waves suitable for auditory stimulation

In a subsequent step, we assessed delta band power and its changes due to PTAS in all 4 types of windows for RS1-3. Additionally, in RS2 and RS3, we probed whether the DBS status (DBS+, on vs. DBS-, off) affected the performance of the PTAS algorithm and the response to PTAS. To address these questions, we first assessed the overall sleep macrostructure across all three Recording Sessions based on the visually scored sleep stages (Supplementary Table SM). The overall sleep efficiency was consistently above 80% and patients spent most of their sleep time in NREM stages 2 and 3. Importantly, no differences were found between the DBS+ and DBS-Sessions, indicating that the DBS status did not influence sleep architecture in the patients.

To assess the number of slow waves suitable for PTAS across the three Recording Sessions, we calculated the mean number of triggers per each type of window: stim ON, stim OFF, sham ON, and sham OFF (Fig. 3). On average in all Recording Sessions, there were 5.0 triggers in stim ON and sham ON windows, and 4.3 and 4.5 in stim OFF and sham OFF, respectively. To estimate the effect of *DBS status* (+ or -), *Condition* (stim vs. sham), and *Window* (ON vs. OFF) on the mean number of triggers, a three-way ANOVA was performed. We observed a significant main effect of *Window*, indicating a lower mean number of triggers in OFF windows. Furthermore, there was a significant interaction of *Window*Condition*, which is explained by a lower mean number of triggers in stim as compared to sham in OFF windows [F(1,6)=11.2; p=0.02]. *DBS status* did not affect the mean number of triggers per window.

**Figure 3.**
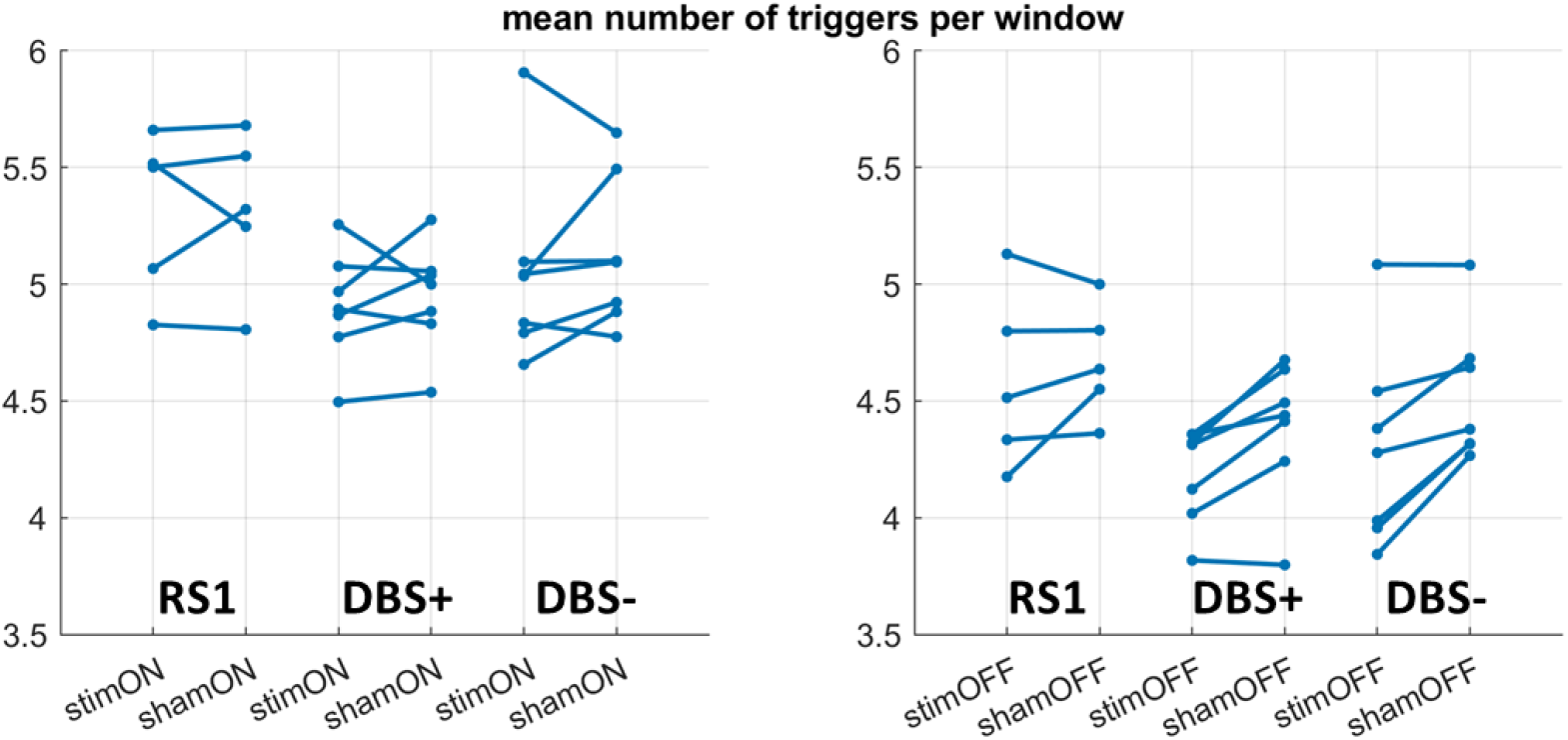
Mean number of triggers per window. (slow waves detected in Fpz fulfilling criteria for auditory stimulation) during Recording Session 1, DBS+, and DBS-Sessions.

### 3.2. EEG power spectral density

The spectral analysis (Fig. 4) of the RS1 data revealed a broadband increase in power of the Fpz signal, including the delta frequency band (0.5-4 Hz) in stim ON windows as compared to sham ON (and the same contrast for OFF windows). The power of STN-LFP was also modulated by PTAS, with statistically significant changes in the low delta (∼0.5-2 Hz), high theta (∼6-8 Hz), and sigma (∼11-18 Hz) ranges.

**Figure 4.**
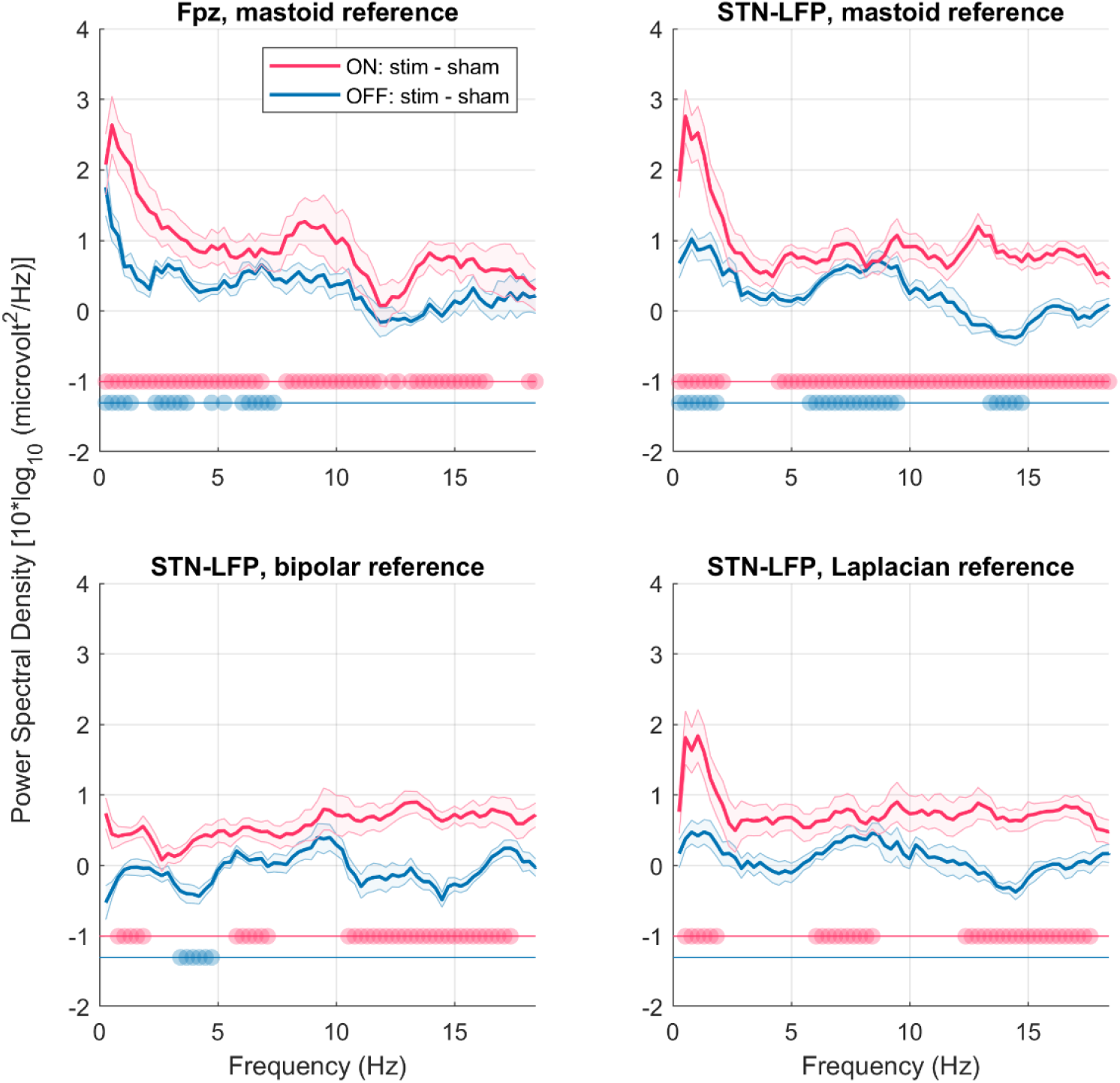
Difference in power spectral density for Recoding Session 1 (different reference montages) between stimulation and sham conditions. Data is presented for detection electrode Fpz (averaged mastoid reference, N=5, mean±CI) and STN-LFP (average mastoid, bipolar, and Laplacian reference, N=10, mean±CI). Data is presented for two contrasts revealing the auditory stimulation effect: stim ON minus sham ON (pink line) and stim OFF minus sham OFF (blue line). Scatter dots indicate p<0.05 for the corresponding comparison (not corrected for Fpz and cluster-corrected for STN-LFP).

The spectral analysis of the Fpz signal for RS2 and RS3 (DBS+ and DBS-) has shown that irrespective of the DBS status, there were significant differences between stim ON and sham ON windows in the broad frequency range, similar to RS1 (Fig. 5). Notably, there was a significant enhancement in OFF windows, which could be explained by a carry-over effect.

**Figure 5.**
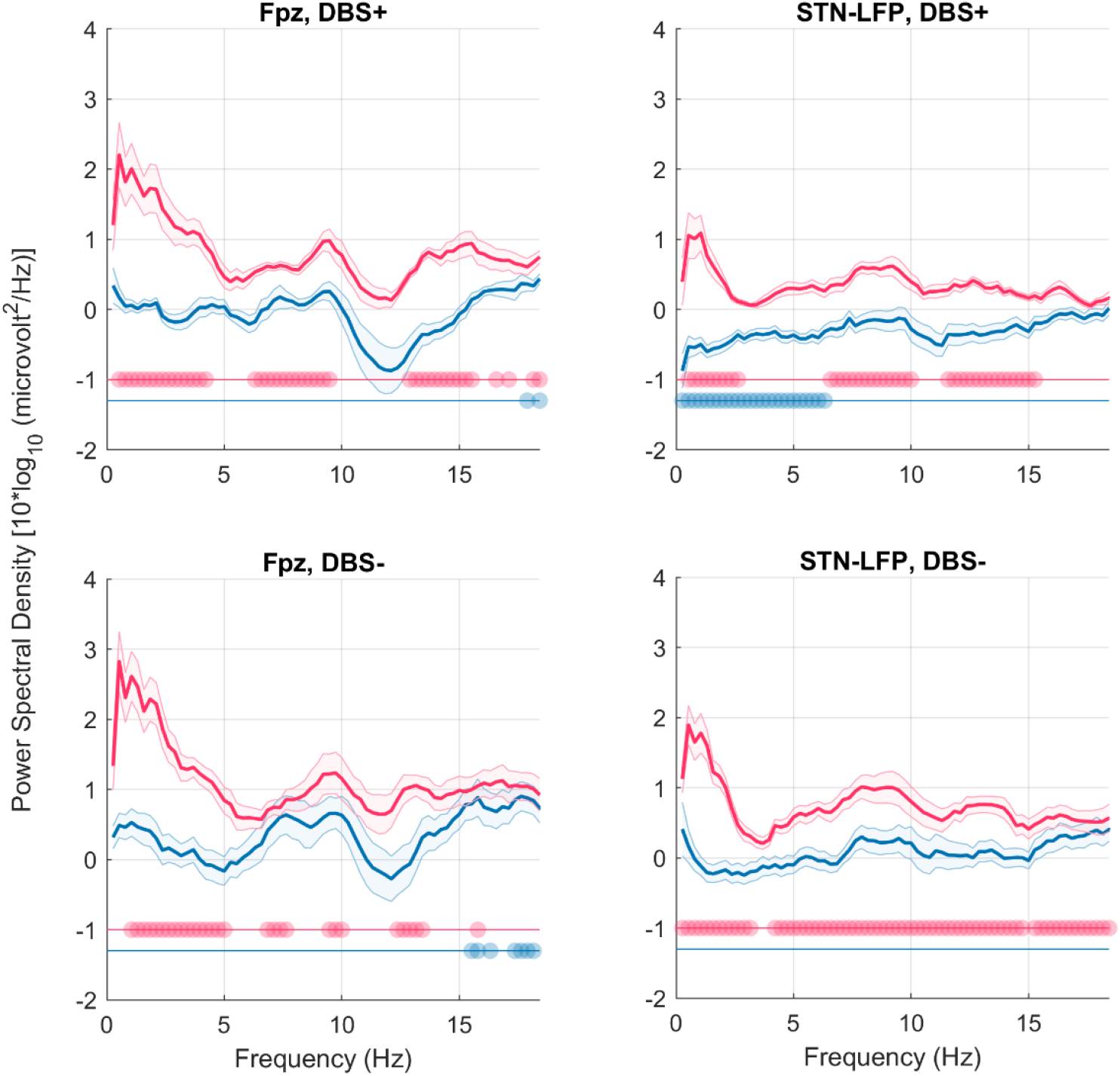
Difference in power spectral density for DBS+ (upper row) and DBS- (bottom row) Recording Sessions between stimulation and sham conditions. Data is presented for detection electrode Fpz (N=5, mean±CI) and STN-LFP (N=10, mean±CI). Data is presented for two contrasts revealing the auditory stimulation effect: stim ON minus sham ON (pink line) and stim OFF minus sham OFF (blue line). Scatter dots indicate p<0.05 for the corresponding comparison (not corrected for Fpz and cluster-corrected for STN-LFP). Of note, there was a hardware highpass filter at 1 Hz applied to the Percept device recording.

To evaluate the effect of *DBS status* (+ or -), *Condition* (stim vs. sham), and *Window* (ON vs. OFF) on the low-delta STN-LFP power (1-2 Hz) we performed three-way ANOVA on power values. The main effects of *Window* and *Condition* were significant. There was a significant interaction of *Window*Condition*, which is explained by an increase of power in ON as compared to OFF only in stim [F(1,9)=17.7; p=0.002]. Another significant interaction was found for *DBS status*Condition*, due to the increase in power in stim condition only for DBS-[F(1,9)=8.9; p=0.02], but the main effect of *DBS status* was not significant.

### 3.3. Coherence between the surface EEG and LFP

In the next step, we investigated how coherent the surface frontal EEG and STN-LFP signals are within four types of PTAS windows. We explored how three reference montages for RS 1 are affecting the measure of magnitude-squared coherence between Fpz and STN-LFP (Fig. 6). The coherence was most pronounced in the low delta range and was statistically significant from zero in four types of PTAS windows (FDR corrected p<0.02, coherence averaged from 0.5 to 2 Hz) for all reference montages. PTAS significantly increased the coherence in the delta range (stim ON vs. sham ON comparison, Fig. 6).

**Figure 6.**
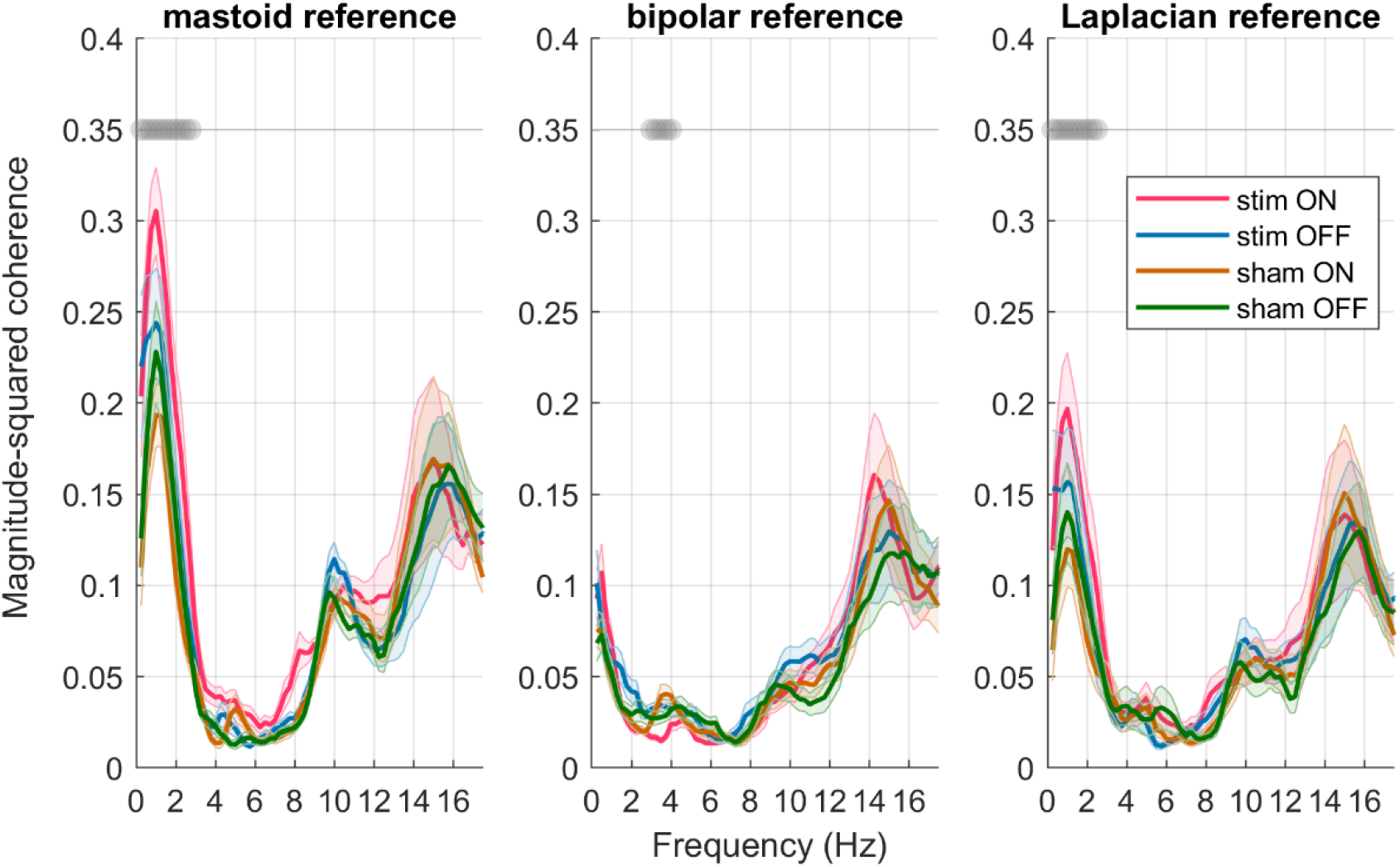
Coherence between Fpz and STN-LFP for the Recording Session 1. Coherence was calculated for the mastoid-referenced Fpz signal and STN-LFP referenced in three different ways (mastoid, bipolar, and Laplacian reference montages, N=10, mean±CI). Four types of windows for auditory stimulation are color-coded. Gray dots indicate p<0.05 for stim ON vs. sham ON comparison, cluster corrected.

Coherence between surface EEG and STN-LFP in DBS+ and DBS-Sessions (Fig. 7) was also significantly different from zero (FDR corrected p<0.01, 0.5-2 Hz), but overall rather low. To assess the effect of *DBS status* (+ or -), *Condition* (stim vs. sham), and *Window* (ON vs. OFF) on the low-delta coherence we performed three-way ANOVA. The main effects of factors *Window* and *Condition* were significant, and there was an interaction of *Window*Condition*, which is explained by a stronger coherence in ON as compared to OFF only in stim [F(1,9)=8.8; p=0.02]. *DBS status* did not affect coherence between surface EEG and STN-LFP in DBS+ and DBS-Sessions.

**Figure 7.**
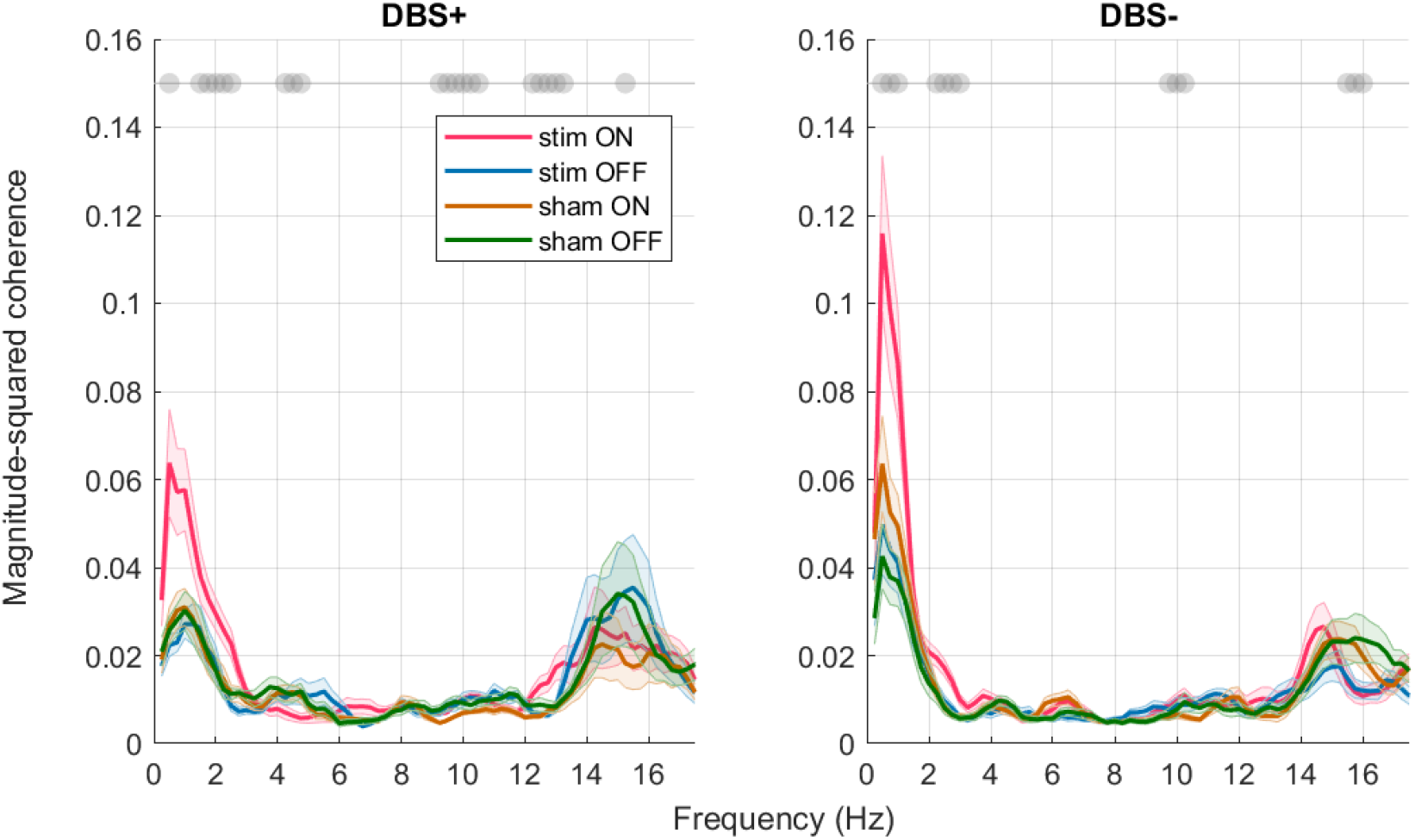
Coherence between Fpz and STN-LFP signals for Recording Sessions 2 and 3 (DBS+ and DBS-). Coherence was calculated for the mastoid-referenced Fpz signal and bipolar-referenced STN-LFP (N=10, mean±CI). Four types of windows for auditory stimulation are color-coded. Gray dots indicate p<0.05 for stim ON vs. sham ON comparison, not cluster corrected.

## 4. Discussion

### 4.1. Coherent delta activity of the surface EEG and STN-LFP signal

In this study, we collected a unique dataset of simultaneous surface EEG and STN-LFP recordings during full-night sleep in PD patients with implanted STN-DBS leads. The recordings were performed for three nights: immediately after the leads were implanted in the STN (RS1) and during the rehabilitation period under DBS+ (on) and DBS-(off) conditions (RS2 and RS3). To address our main research question on whether the STN-LFP signal could be used for PTAS, we first tested the detectability of cortical slow waves in the STN-LFP across all Recording Sessions. Prior research in animals has shown that cortically generated sleep slow waves (Nir et al., 2011; Krone et al., 2021) are reflected in subcortical structures. The discharge burst of the STN neurons was found to be temporally aligned with the rising phase of cortical slow waves (Magill et al., 2000). Furthermore, in the same study it was found that when the cortex was pharmacologically inactivated, the temporal pattern of bursting STN activity was lost. Authors have concluded that during NREM, delta-range oscillatory activity of the cortex is transmitted to the STN-globus pallidus network. Recently, it was found that slow-wave sleep can be identified using STN recordings only (Smyth et al., 2023). In our study, by performing the offline analysis of slow waves in STN-LFP and the ERP analysis, we confirm that slow waves detected in the cortex can also be captured in the STN. Supporting these findings, we observed a similar increase in delta-range activity in both the frontal surface channel and STN-LFP during PTAS. Notably, this delta-range activity was coherent between the surface and STN-LFP signals irrespective of the time after the surgery and DBS status. Although the temporal synchronization of the surface EEG and STN-LFP was performed manually in RS2 and RS3, and therefore, less precise, the coherence between the two signals was still present. DBS status did not affect the coherence, indicating that it is feasible to detect surface slow waves in the STN-LFP during NREM sleep without switching off DBS.

### 4.2. Effect of phase-targeted auditory stimulation on the surface EEG and STN-LFP signal

When evaluating the impact of PTAS on spectral power, we found a consistent broadband enhancement of power across all Recording Sessions. Importantly, the effect was always present in the low delta range, even in the case of STN-LFP recordings performed with the Percept device, where a 1 Hz hardware filter was applied. Thus, we can conclude that PTAS effectively boosted slow waves irrespective of DBS status, and remained detectable even when hardware limitations were present. Of note, the analysis of the mean number of triggers per window has shown a decrease in the stim OFF windows as compared to sham OFF. This observation could be explained by the refractory period, following a train of slow waves evoked by the PTAS (Bernardi et al., 2018), effectively resulting in a “redistribution” of slow waves towards the ON windows.

PTAS also increased coherence in the delta range, which might be related to the simultaneous propagation of the evoked slow waves both through cortical areas and the STN. Alternatively, as coherence does not strictly reflect only the phase relationships but can be also influenced by the amplitude covariation, the observed PTAS-driven increase of coherence can be related to the change in power of both signals (Lachaux et al., 1999).

The exploratory ERP analysis confirmed that the PTAS effect can be assessed from surface EEG and STN-LFP signals. Detecting ERPs in the intracranial signal holds significant importance, as it can indicate whether the volume of the presented stimuli is sufficient to evoke sensory system response, or if it falls below the perception threshold. Consequently, the presence of distinguishable ERP in STN-LFP in the future can aid in tuning the PTAS parameters for individual patients.

### 4.3. Reference montage for STN-LFP signal in Recording Session 1

When comparing different reference methods for STN-LFP of RS1, we found that using a mastoid reference for STN-LFP resulted in the greatest similarity to the surface-EEG signal. However, it would be impossible to use such a reference method for the solution based solely on STN-implanted leads. The two within-STN reference montages that we tested are the bipolar montage (the one that is used for the recording in the BrainSense mode of the Percept device) and Laplacian montage, highlighting the focal intra-SNT activity by subtracting from it the signals of neighboring channels. While the results were rather similar for these two referencing montages, the Laplacian montage has shown a more pronounced delta-range peak in spectral power and coherence analysis. The observation of the higher coherence in the Laplacian montage agrees with the offline wave analysis. The better outcome for the Laplacian montage as compared to bipolar can be explained by the lack of consistency in the location of the two electrodes used for the bipolar montage recordings (for details see methods, Supplementary Table CL). The active electrode, whose signal was accentuated with Laplacian montage, was always located within STN. The two recording electrodes used for the bipolar recordings, due to the small size of STN can be located within STN, substantia nigra, or surrounding white matter. This configuration, however, allows cancellation of the DBS artifact, and bipolar recording is the only option to register STN activity when DBS is switched on.

To sum up the reference schemes analysis findings, an internal reference located in the white matter would be an optimal solution for the detection of slow waves in the STN signal. For example, an additional contact could be located more dorsally along the lead in the subcortical frontal white matter. Such reference contact would provide an excellent signal-to-noise ratio and would allow capturing large slow waves in the STN-LFP signal, which are lost in bipolar referencing.

### 4.4. Study limitations

There are several limitations of this study that we would like to address. This work relies on data from a small cohort of patients, and in future studies, it would be important to establish how the individual diagnosis specifics can impact the detectability of slow waves in STN. The patient-specific DBS lead location for the bipolar recordings RS2 and RS3 is another factor that makes the interpretation of the results more difficult. Furthermore, when performing night sleep recordings, we encountered several hardware limitations of the Percept device, such as embedded 1 Hz filter, 4-h limits of the recording duration, and challenges in synchronizing the Percept device recording with the external EEG amplifier.

### 4.5. Conclusion and outlook

This work provides the first evidence that the detection of NREM slow waves in STN-LFP signal is possible, which might enable the implementation of PTAS protocol based solely on STN-LFP. Furthermore, we could demonstrate that the effects of PTAS can be detected in the STN-LFP signal, which opens the possibility to estimate the efficiency of PTAS to boost slow waves without surface recordings. These findings offer new avenues for PTAS implementation to improve sleep in patients with PD.

## Data Availability

All data produced in the present study are available upon reasonable request to the authors

https://drive.google.com/file/d/1aRPlhPQgRgvtkIQwNITdolavyOolkTTa/view?usp=sharing

## Abbreviations

CT: computed tomography
DBS: deep brain stimulation
EEG: electroencephalography
EOG: electrooculography
ERP: event-related potentials
FIR: finite impulse response
LFP: local field potential
MDS-UPDRS: Movement Disorder Society-Sponsored Revision of the Unified Parkinson’s Disease Rating Scale
MRI: magnetic resonance imaging
NREM: non-rapid eye movement sleep
PD: Parkinson’s disease
PLL: phase-locked loop
PSG: polysomnography
PTAS: phase-targeted auditory stimulation
REM: rapid eye movement sleep
RS: recording session
STN: local field potential
SWS: slow-wave sleep

## 5. Author contributions

Elena Krugliakova: investigation, data curation, formal analysis, methodology, software, validation, visualization, writing - original draft, writing - review & editing;

Artyom Karpovich: project administration, investigation, formal analysis, writing - review & editing; Maria Jacomet: investigation, formal analysis, writing - review & editing;

Stephanie Huwiler and Caroline Lustenberger: methodology, software, writing - review & editing; Lennart Stieglitz: resources, data curation, formal analysis, writing - review & editing;

Lukas Imbach, Bartosz Bujan, Piotr Jedrysiak: methodology, resources, writing - review & editing;

Christian R. Baumann: conceptualization, funding acquisition, resources, supervision, writing - review & editing;

Sara Fattinger: conceptualization, funding acquisition, supervision, project administration, investigation, formal analysis, writing - original draft, writing - review & editing.

## 6. Acknowledgements

The authors would like to thank the patients; staff of the University Hospital of Zurich and Clinic Lengg (Zurich, Switzerland especially Dr. med. Fabian Büchele, Dr. med. Evdokia Efthymiou, Mechtild Uhl, Lea Mannale, and Adnan Sopi); Prof. Reto Huber, Prof. Peter Achermann, and Simone Accascina for their advice on experimental setup and data analysis; Natalie Birnbaum for her contribution in the data collection process.

## 7. Declaration of generative AI and AI-assisted technologies in the writing process

During the preparation of this work the authors used tools ChatGPT 3.5 and Grammarly to check for spelling and grammar mistakes in the text body. After using this tool, the authors considered some of the suggestions that were made and implemented them. Authors take full responsibility for the content of the publication.

## References

Baumann-Vogel, H., Imbach, L. L., Sürücü, O., Stieglitz, L., Waldvogel, D., Baumann, C. R., et al. (2017). The Impact of Subthalamic Deep Brain Stimulation on Sleep-Wake Behavior: A Prospective Electrophysiological Study in 50 Parkinson Patients. Sleep 40. doi:10.1093/sleep/zsx033.

Baumann-Vogel, H., Valko, P. O., Eisele, G., and Baumann, C. R. (2015). Impulse control disorders in Parkinson’s disease: don’t set your mind at rest by self-assessments. Eur. J. Neurol. 22, 603–609. doi:10.1111/ene.12646.

Benabid, A. L., Pollak, P., Gervason, C., Hoffmann, D., Gao, D. M., Hommel, M., et al. (1991). Long-term suppression of tremor by chronic stimulation of the ventral intermediate thalamic nucleus. Lancet (London, England) 337, 403–6. doi:10.1016/0140-6736(91)91175-t.

Benabid, A. L., Pollak, P., Gross, C., Hoffmann, D., Benazzouz, A., Gao, D. M., et al. (1994). Acute and Long-Term Effects of Subthalamic Nucleus Stimulation in Parkinson’s Disease. Stereotact. Funct. Neurosurg. 62, 76–84. doi:10.1159/000098600.

Bernardi, G., Siclari, F., Handjaras, G., Riedner, B. A., and Tononi, G. (2018). Local and widespread slow waves in stable NREM sleep: evidence for distinct regulation mechanisms. Front. Hum. Neurosci. 12, 248. doi:10.3389/fnhum.2018.00248.

Büchele, F., Hackius, M., Schreglmann, S. R., Omlor, W., Werth, E., Maric, A., et al. (2018). Sodium oxybate for excessive daytime sleepiness and sleep disturbance in Parkinson disease: A randomized clinical trial. JAMA Neurol. 75, 114. doi:10.1001/jamaneurol.2017.3171.

Chiu, N. T., Huwiler, S., Ferster, M. L., Karlen, W., Wu, H. T., and Lustenberger, C. (2022). Get rid of the beat in mobile EEG applications: A framework towards automated cardiogenic artifact detection and removal in single-channel EEG. Biomed. Signal Process. Control 72, 103220. doi:10.1016/j.bspc.2021.103220.

De Cock, V. C., Vidailhet, M., and Arnulf, I. (2008). Sleep disturbances in patients with parkinsonism. Nat. Clin. Pract. Neurol. 4, 254–266. doi:10.1038/ncpneuro0775.

Fattinger, S., de Beukelaar, T. T., Ruddy, K. L., Volk, C., Heyse, N. C., Herbst, J. A., et al. (2017). Deep sleep maintains learning efficiency of the human brain. Nat. Commun. 8, 15405. doi:10.1038/ncomms15405.

Feigin, V. L., Abajobir, A. A., Abate, K. H., Abd-Allah, F., Abdulle, A. M., Abera, S. F., et al. (2017). Global, regional, and national burden of neurological disorders during 1990–2015: a systematic analysis for the Global Burden of Disease Study 2015. Lancet Neurol. 16, 877–897. doi:10.1016/S1474-4422(17)30299-5.

Gerstner, J. R., Perron, I. J., and Pack, A. I. (2012). The nexus of Aβ, aging, and sleep. Sci. Transl. Med. 4, 2–5. doi:10.1126/scitranslmed.3004815.

Geva-Sagiv, M., Mankin, E. A., Eliashiv, D., Epstein, S., Cherry, N., Kalender, G., et al. (2023). Augmenting hippocampal–prefrontal neuronal synchrony during sleep enhances memory consolidation in humans. Nat. Neurosci. 26, 1100–1110. doi:10.1038/s41593-023-01324-5.

Hablitz, L. M., Vinitsky, H. S., Sun, Q., Stæger, F. F., Sigurdsson, B., Mortensen, K. N., et al. (2019). Increased glymphatic influx is correlated with high EEG delta power and low heart rate in mice under anesthesia. Sci. Adv. 5. doi:10.1126/sciadv.aav5447.

Huwiler, S., Carro Dominguez, M., Huwyler, S., Kiener, L., Stich, F. M., Sala, R., et al. (2022). Effects of auditory sleep modulation approaches on brain oscillatory and cardiovascular dynamics. Sleep 45, 1–18. doi:10.1093/sleep/zsac155.

Krone, L. B., Yamagata, T., Blanco-Duque, C., Guillaumin, M. C. C., Kahn, M. C., van der Vinne, V., et al. (2021). A role for the cortex in sleep–wake regulation. Nat. Neurosci. 24, 1210–1215. doi:10.1038/s41593-021-00894-6.

Krugliakova, E., Skorucak, J., Sousouri, G., Leach, S., Snipes, S., Ferster, M. L., et al. (2022). Boosting Recovery During Sleep by Means of Auditory Stimulation. Front. Neurosci. 16, 1–13. doi:10.3389/fnins.2022.755958.

Lachaux, J. P., Rodriguez, E., Martinerie, J., and Varela, F. J. (1999). Measuring phase synchrony in brain signals. Hum. Brain Mapp. 8, 194–208. doi:10.1002/(SICI)1097-0193(1999)8:4<194::AID-HBM4>3.0.CO;2-C.

Lhommée, E., Klinger, H., Thobois, S., Schmitt, E., Ardouin, C., Bichon, A., et al. (2012). Subthalamic stimulation in Parkinson’s disease: restoring the balance of motivated behaviours. Brain 135, 1463–77. doi:10.1093/brain/aws078.

Lhommée, E., Wojtecki, L., Czernecki, V., Witt, K., Maier, F., Tonder, L., et al. (2018). Behavioural outcomes of subthalamic stimulation and medical therapy versus medical therapy alone for Parkinson’s disease with early motor complications (EARLYSTIM trial): secondary analysis of an open-label randomised trial. Lancet Neurol. 17, 223–231. doi:10.1016/S1474-4422(18)30035-8.

Lustenberger, C., Ferster, M. L., Huwiler, S., Brogli, L., Werth, E., Huber, R., et al. (2022). Auditory deep sleep stimulation in older adults at home: a randomized crossover trial. Commun. Med. 2. doi:10.1038/s43856-022-00096-6.

Magill, P. J., Bolam, J. P., and Bevan, M. D. (2000). Relationship of activity in the subthalamic nucleus-globus pallidus network to cortical electroencephalogram. J. Neurosci. 20, 820–833. doi:10.1523/jneurosci.20-02-00820.2000.

Morawska, M. M., Moreira, C. G., Ginde, V. R., Valko, P. O., Weiss, T., Büchele, F., et al. (2021). Slow-wave sleep affects synucleinopathy and regulates proteostatic processes in mouse models of Parkinson’s disease. Sci. Transl. Med. 13. doi:10.1126/scitranslmed.abe7099.

Ngo, H. V. V., Martinetz, T., Born, J., and Mölle, M. (2013). Auditory closed-loop stimulation of the sleep slow oscillation enhances memory. Neuron 78, 545–553. doi:10.1016/j.neuron.2013.03.006.

Nir, Y., Staba, R. J., Andrillon, T., Vyazovskiy, V. V., Cirelli, C., Fried, I., et al. (2011). Regional slow waves and spindles in human sleep. Neuron 70, 153–169. doi:10.1016/j.neuron.2011.02.043.

Noble, W., and Spires-Jones, T. L. (2019). Sleep well to slow Alzheimer’s progression? Science (80-.). 363, 813–814. Available at: http://www.embase.com/search/results?subaction=viewrecord&from=export&id=L2001626535*0A z10.1126/science.aaw5583.

Oostenveld, R., Fries, P., Maris, E., Schoffelen, J.-M., Oostenveld, R., Fries, P., et al. (2011). FieldTrip: open source software for advanced analysis of MEG, EEG, and invasive electrophysiological data. Comput. Intell. Neurosci. 2011, e156869. doi:10.1155/2011/156869.

Papalambros, N. A., Weintraub, S., Chen, T., Grimaldi, D., Santostasi, G., Paller, K. A., et al. (2019). Acoustic enhancement of sleep slow oscillations in mild cognitive impairment. Ann. Clin. Transl. Neurol. doi:10.1002/acn3.796.

Prehn-Kristensen, A., Ngo, H. V. V., Lentfer, L., Berghäuser, J., Brandes, L., Schulze, L., et al. (2020). Acoustic closed-loop stimulation during sleep improves consolidation of reward-related memory information in healthy children but not in children with attention-deficit hyperactivity disorder. Sleep 43, zsaa017. doi:10.1093/sleep/zsaa017.

Pringsheim, T., Jette, N., Frolkis, A., and Steeves, T. D. L. (2014). The prevalence of Parkinson’s disease: A systematic review and meta-analysis. Mov. Disord. 29, 1583–1590. doi:10.1002/mds.25945.

Roh, J. H., Huang, Y., Bero, A. W., Kasten, T., Stewart, F. R., Bateman, R. J., et al. (2012). Disruption of the sleep-wake cycle and diurnal fluctuation of amyloid-β in mice with Alzheimer’s disease pathology. Sci. Transl. Med. 4. doi:10.1126/scitranslmed.3004291.

Schreiner, S. J., Imbach, L. L., Werth, E., Poryazova, R., Baumann-Vogel, H., Valko, P. O., et al. (2019). Slow-wave sleep and motor progression in Parkinson disease. Ann. Neurol. 85, 765–770. doi:10.1002/ana.25459.

Sharma, M., Naik, V., and Deogaonkar, M. (2016). Emerging applications of deep brain stimulation. J. Neurosurg. Sci. 60, 242–55.

Siegfried, J., and Lippitz, B. (1994). Bilateral chronic electrostimulation of ventroposterolateral pallidum: a new therapeutic approach for alleviating all parkinsonian symptoms. Neurosurgery 35, 1126–9; discussion 1129-30. doi:10.1227/00006123-199412000-00016.

Smyth, C., Anjum, M. F., Ravi, S., Denison, T., Starr, P., and Little, S. (2023). Adaptive Deep Brain Stimulation for sleep stage targeting in Parkinson’s disease. Brain Stimul. 16, 1292–1296. doi:10.1016/j.brs.2023.08.006.

Sousouri, G., Baumann, C. R., Imbach, L. L., Huber, R., and Werth, E. (2021). Sleep electroencephalographic asymmetry in Parkinson’s disease patients before and after deep brain stimulation. Clin. Neurophysiol. doi:10.1016/j.clinph.2020.12.027.

Sürücü, O., Baumann-Vogel, H., Uhl, M., Imbach, L. L., and Baumann, C. R. (2013). Subthalamic deep brain stimulation versus best medical therapy for l-dopa responsive pain in Parkinson’s disease. Pain 154, 1477–1479. doi:10.1016/j.pain.2013.03.008.

Tononi, G., and Cirelli, C. (2014). Sleep and the price of plasticity: from synaptic and cellular homeostasis to memory consolidation and integration. Neuron 81, 12–34. doi:S0896-6273(13)01186-0 [pii]10.1016/j.neuron.2013.12.025.

Vyazovskiy, V. V, and Harris, K. D. (2013). Sleep and the single neuron: the role of global slow oscillations in individual cell rest. Nat Rev Neurosci. doi:nrn3494 [pii]10.1038/nrn3494.

Xie, L., Kang, H., Xu, Q., Chen, M. J., Liao, Y., Thiyagarajan, M., et al. (2013). Sleep drives metabolite clearance from the adult brain. Science (80-.). 342, 373–377. doi:10.1126/science.1241224.

