## Supplementary information for "Exploring the local field potential signal from the subthalamic nucleus for phase-targeted auditory stimulation in Parkinson’s disease"

### 1. Supplementary methods

#### 1.1. Detailed information on participants recruitment and screening

A total of 15 patients diagnosed with PD (mild to moderate disease severity, Hoehn-Yahr stages ll-lll) undergoing bilateral STN-DBS implantation at University Hospital Zurich were included in the study. Initial preoperative inclusion criteria were the following: age above 18, negative pregnancy test, and sufficient German language skills to follow study procedures. Exclusion criteria were regular use of benzodiazepines, use of melatonin less than 1 day prior to the recording session, hearing deficiency, resulting in an inability to hear the auditory stimuli during sleep (based on results of standard pure-tone threshold audiometry). Prior to the first surgery during a polysomnographic (PSG) recording in the framework of a standardized clinical pre-DBS work-up, a Screening night was performed to assess sleep-related physiological exclusion criteria: the presence of clinical moderate to severe sleep-wake disorders (such as restless leg syndrome, periodic limb movement disorder, and apnoea). Screening night EEG data was also used to test the optimal phase-targeted auditory stimulation (PTAS) parameters. In three patients PSG night recording was combined with PTAS (applied based on slow waves detected in the EEG signal), in order to perform the screening for susceptibility and tolerance to PTAS. In eight patients PTAS was not applied during the PSG recording, but the collected EEG data was used to perform simulations of PTAS algorithm performance. Both screening PTAS and simulated PTAS outcomes were used to ensure the selection of the optimal parameters for PTAS algorithm non-rapid eye movement sleep (NREM) sleep detection and maximize the number of stimuli applied for each patient during the recording sessions.

Four participants dropped out before the start of the data recording. 11 patients (5 females, age range 51-75 years old) have participated in the data recordings. Only 9 patients had sleep efficiency more than 70% either during Recording Session 1 (RS1) or both Recording Sessions 2 (RS2) and 3 (RS3), and were included in the analysis (see Supplementary Table PC for the demographic information).

In 5 out of these 9 participants, it was possible to record data during RS1: Only in patients with full-ring leads the implantation of the leads and Medtronic device was performed with externalization of contacts in between surgeries (for more details, see Supplementary methods, Information on surgical procedure section). Moreover, in 7 patients both DBS+ and DBS- night were successfully completed, in 2 LFP data was lost during one of the recording sessions due to technical issues. As a result, for the DBS+ and DBS- nights, we could include all 7 patients in the analysis not related to the LFP signal processing (sleep structure, number of triggers) and 5 patients in the LFP data analysis (see Supplementary Table AT).

#### 1.2. Information on surgical procedure

Five patients were implanted with Medtronic (Medtronic Neurological Division, Minneapolis, MN, USA) lead model 3389 (addressed as full-ring leads), and six with model B33005 (tripartite directional leads). The implanted Medtronic leads have four cylindrical (full-ring or tripartite) contacts denominated 0–1–2–3 for the left and 8–9–10–11 for the right side beginning from the more ventral contact (Fig. 1, A). Accurate implantation within the STN was intraoperatively verified by micro-electrode recordings (Leadpoint, Medtronic Neurological Division, Minneapolis, MN, USA) in steps of 0.5 mm until the pars reticulata of the substantia nigra (starting 10 mm above the target), as well as clinical response upon intraoperative stimulation and intraoperative computed tomography (CT). The accurate lead placement was verified by comparing the planned and actual leads position in the post-surgery CT scan (Fig. SIL, B, C). In the case of full-ring leads, the internalization and connection of the lead wires to the stimulation device (Percept™ PC, Medtronic Neurological Division, Minneapolis, MN, USA) was performed 3–5 days after the first surgery (two-step surgical approach).

#### 1.3. Phase-targeted auditory stimulation

In brief stimulation algorithm can be described in two parts: stable NREM sleep detection and phase-targeting algorithm. Stable NREM sleep was defined as 10 min of uninterrupted NREM sleep. The NREM sleep was detected, when low delta (2–4 Hz), high delta (3–5 Hz), and high beta (20–30 Hz) power of the Fpz signal (online re-referenced to right mastoid) met the predefined thresholds. A correlation index for two EOG signals was used to identify REM sleep periods: whenever a strong negative correlation was present, NREM sleep was not detected. To target the slow waves of sufficient amplitude, another threshold was used based on the power in the delta frequency range. Additionally, beta power was estimated, and movement detection (sharp large-amplitude artifacts) was used to avoid stimulation during arousals. To deliver stimuli at a specific phase of ongoing slow waves, a first-order phase-locked loop (PLL) was used according to Ferster et al., 2022. Every ON window started after the following criteria were satisfied: target phase of the PLL has been reached, stable NREM sleep, beta power below the threshold, no movement detected, sufficiently large-amplitude slow waves, and low EOG negative correlation index. Those criteria are described in more detail in Lustenberger et al., 2022. Thus, the beginning of the ON window always coincided with the first stimulus presentation. The minimal inter-stimulus interval within the window was 500 ms.

To control for the possible delays in stimuli presentation due to hardware limitations (e.g., the delay introduced by stimulating the laptop's soundcard), the StimTrak module was used (Brain Products GmbH, Gilching, Germany). The average delay was ~130 ms, during the data analysis trigger positions were adjusted.

### 1.4. Synchronization of the surface EEG and STN-LFP in Recording Sessions 2 and 3

To synchronize surface EEG and LFP recording, the electrical artifact induced by the DBS was used to subsequently align LFP and surface EEG offline. In more detail, DBS via the Percept PC can be used to generate a stimulation-induced artifact detectable in the LFP recording, but it can also be picked up by the external EEG amplifier. Because the usual therapeutic DBS stimulation frequency is above the Nyquist frequency (125 Hz) of the used sampling rate (250 Hz), we used an artifact produced at 85 or 90 Hz. Thus, in our case, a sharp change in 85/90 Hz power (DBS was turned "on" and "off" in BrainSense Streaming mode) was registered by both Percept PC and BrainAmp DC amplifier. During offline data processing, we performed wavelet analysis to capture change in the 85/90 Hz frequency band in both signals. LFP recording was aligned to the surface EEG using the resulting timestamps.

### 1.5. Sleep scoring

For sleep scoring, the surface EEG data were consequently high-pass (finite impulse response (FIR) filter, 0.5 Hz) and low-pass filtered (FIR filter, 40 Hz), re-referenced to the earlobes, and downsampled to 128 Hz. Vigilance states (wake, N1, N2, N3, and REM) were visually scored by a sleep expert and verified by another sleep expert (author SF). Scoring was performed using the signal from frontal, central, and occipital electrodes (20-s epochs) based on the American Academy of Sleep Medicine standard criteria (Berry et al., 2015).

### 1.6. Cluster correction of p-values

The cluster correction procedure was used over frequency bins in the spectrogram and coherence plot, over time points in ERPs. As an example, in the case of the spectrogram, first, a paired t-test for the contrast of interest was performed separately for all frequency bins. Next, significant neighboring bins were clustered if they showed the same direction of effect. To assess the statistical significance of each cluster, a cluster-level test statistic was calculated by computing the sum of all t-values in the cluster. The significance of each cluster was estimated by comparing the cluster-level test statistic to a reference permutation distribution derived from the data. The reference distribution was obtained by randomly permuting the data 1,000 times. The cluster p-value was estimated as the proportion of the elements in the reference distribution exceeding the cluster-level test statistic. An analogous procedure was used for the other types of analysis.

### 2. Supplementary tables

**Supplementary Table PC. Patient's characteristics prior to surgery.**

Data are presented only for participants included in the data analysis. Abbreviations: H&Y – Hoehn & Yahr scale, LED – Levodopa Equivalent Dose, y – year

| Patient | Sex | Age [5-year range] | Disease subtype | H&Y | Disease duration [y] | Medication | LED [mg] | UPDRS III before surgery |
|---|---|---|---|---|---|---|---|---|
| 1 | f | 55-60 | mixed | 2 | 7 | Madopar DR | 1200 | 19 |
| 2 | f | 55-60 | rigid-akinetic | 2 | 7 | Stalevo, Rasagilin | 1297 | 21 |
| 3 | f | 55-60 | mixed | 2 | 5 | Madopar DR, Rasagilin | 600 | 19 |
| 4 | f | 65-70 | rigid-akinetic | 2 | 7 | Madopar DR, Apomorphine | 754 | 20 |
| 5 | f | 70-75 | tremor dominant | 2 | 6 | Rasagilin, Pramipexol | 300 | 35 |
| 6 | m | 60-65 | rigid-akinetic | 2 | 11 | Stalevo, Rasagilin, Madopar Liq | 1000 | 14 |
| 7 | m | 75-80 | tremor dominant | 2 | 8 | Madopar, Pramipexol | 800 | 44 |
| 8 | m | 50-55 | rigid-akinetic | 2 | 8 | Stalevo, Rasagilin, Amantadin | 1200 | 19 |
| 9 | m | 75-80 | rigid-akinetic | 3 | 11 | Ropinirol, Madopar DR, Madopar | 1250 | 27 |

**Supplementary Table AT. Collected data, and stimulation parameters.**

Data are presented only for participants included in the data analysis. Patient 8, marked with the * sign, had developed a cardiac artifact in STN-LFP, the data was cleaned using the brMEGA algorithm.

| Patient | Type of electrodes | SA and AS analysis | RS1 EEG LFP analysis | RS2+3 LFP EEG analysis | Stimulation contacts (left/right) | Stimulation frequency (Hz) | Voltage (V, left/right) |
|---|---|---|---|---|---|---|---|
| 1 | full ring | yes | yes | yes | C2/C10 | 130 | 0.7/0.7 |
| 2 | full-ring | yes | no | no | C2/C10 | - | - |
| 3 | tripartite | - | yes | no | C2/C10 | 145 | 0.5/1 |
| 4 | full-ring | yes | yes | no | C2/C10 | 130 | 1.4/1 |
| 5 | full-ring | yes | no | no | C2/C10 | - | - |
| 6 | tripartite | - | yes | yes | C2/C10 | 145 | 0.7/0.7 |
| 7 | tripartite | - | yes | yes | C2/C10 | 145 | 0.8/1 |
| 8* | full-ring | yes | yes | yes | C2/C9 | 145 | 0.9/0.9 |
| 9 | tripartite | - | yes | yes | C2/C10 | 145 | 0.8/0.8 |

**Supplementary Table CL. Location of DBS lead contacts.**

DBS lead locations were semi-automatically defined using Medtronic SureTune software (Medtronic, Minneapolis, MN). Bold font is used to indicate the DBS stimulation contact. The blue background colour indicates more ventral contacts (C0 and C1 on the left and C8 and C9 on the right), and orange – more dorsal (C2 and C3 on the left and C10 and C11 on the right). For the bipolar referencing during Recording sessions 2 and 3 (two contacts neighbouring to the stimulation contact), the signal of more dorsal contact was subtracted from more ventral. Abbreviations: SN – substantia nigra, STN – subthalamic nucleus.

| Patient | left hemisphere | | | | right hemisphere | | | |
|---|---|---|---|---|---|---|---|---|
| | C0 | C1 | C2 | C3 | C8 | C9 | C10 | C11 |
| 1 | SN | SN | **the lower border of STN** | STN | between STN and SN | STN bottom | **STN up** | upper border of STN |
| 2 | SN | between STN and SN | **STN bottom** | STN up | SN | between STN and SN | **STN bottom** | STN up |
| 3 | SN | between STN and SN | **STN** | upper border of STN | SN | STN bottom | **STN up** | upper border of STN |
| 4 | SN | between STN and SN | **STN bottom** | STN up | between STN and SN | STN bottom | **STN up** | upper border of STN |
| 5 | SN | STN bottom | **STN up** | upper border of STN | SN | between STN and SN | **STN bottom** | STN up |
| 6 | between STN and SN | STN bottom | **STN up** | upper border of STN | lower border of STN | STN bottom | **STN up** | outside STN |
| 7 | between STN and SN | STN middle | **STN up** | upper border of STN | SN | lower border of STN | **STN mid** | upper border of STN |
| 8 | SN | lower border of STN | **STN bottom** | STN up | SN | **lower border of STN** | STN | STN up |
| 9 | SN | between STN and SN | **STN bottom** | STN up | between STN and SN | STN bottom | **STN up** | upper border of STN |

**Supplementary Table SM. Sleep macrostructure.**

Sleep parameters were calculated from the visual scoring of sleep stages (mean±std, N=5 for the Recording Session 1 (RS1) and N=7 for DBS+ and DBS- sessions). Sleep efficiency was calculated by dividing total sleep time with total time in bed. Wake after sleep onset is expressed as the percentage of total time in bed. Sleep stages (NREM sleep, stage N1, stage N2, stage N3, and REM sleep) are expressed as a percentage of total sleep time. There was no statistical difference between the sleep architecture during DBS+ and DBS- sessions (p-values from paired Student's t-test are shown in the last column).

| | **RS1** | **DBS+** | **DBS-** | **p-value** |
|---|---|---|---|---|
| Total time in bed [h] | 7.0 ± 2.5 | 8.2 ± 0.4 | 8.0 ± 0.4 | 0.5 |
| Total sleep time [h] | 6.0 ± 2.5 | 6.9 ± 0.5 | 6.5 ± 0.8 | 0.4 |
| Sleep efficiency [%] | 83.1 ± 9.8 | 84.2 ± 7.3 | 81.6 ± 9.5 | 0.6 |
| Sleep latency [min] | 10.8 ± 7.5 | 17.5 ± 15.7 | 19.4 ± 12.8 | 0.8 |
| Wake after sleep onset [%] | 12.6 ± 6.1 | 11.6 ± 7.0 | 13.7 ± 8.6 | 0.7 |
| NREM sleep [%] | 90.5 ± 7.3 | 81.3 ± 9.6 | 79.9 ± 4.9 | 0.7 |
| Stage N1 [%] | 9.4 ± 4.5 | 7.0 ± 4.8 | 8.4 ± 3.5 | 0.5 |
| Stage N2 [%] | 64.0 ± 4.1 | 53.4 ± 13.6 | 48.7 ± 8.3 | 0.5 |
| Stage N3 [%] | 17.1 ± 11.4 | 21.0 ± 10.8 | 22.7 ± 7.5 | 0.7 |
| REM sleep [%] | 9.5 ± 7.3 | 18.7 ± 9.6 | 20.1 ± 4.9 | 0.7 |

### 3. Supplementary results

#### 3.1. Example of referencing to cardiogram electrode

To address the question whether mastoid electrodes capture slow-wave activity, and thus could influence the amplitude of slow waves observed in STN-LFP data when referenced to mastoid in Recording Session 1, we conducted an exploratory analysis in one patient. First, we detected slow waves in the FPz electrode referenced to the joint mastoid (same as for Fig. 2). These time-points were then used for the time-locked analysis of data filtered in low delta band. Next, we re-referenced both FPz and STN-LFP to either A1 or ECG1 (an electrode located on the participant's chest). Comparing the resulting waveforms revealed that the A1 reference was nearly equivalent to the ECG1 reference. This finding suggests that the A1 signal does not significantly influence the slow waves observed in the LFP signal during the Recording Session 1.

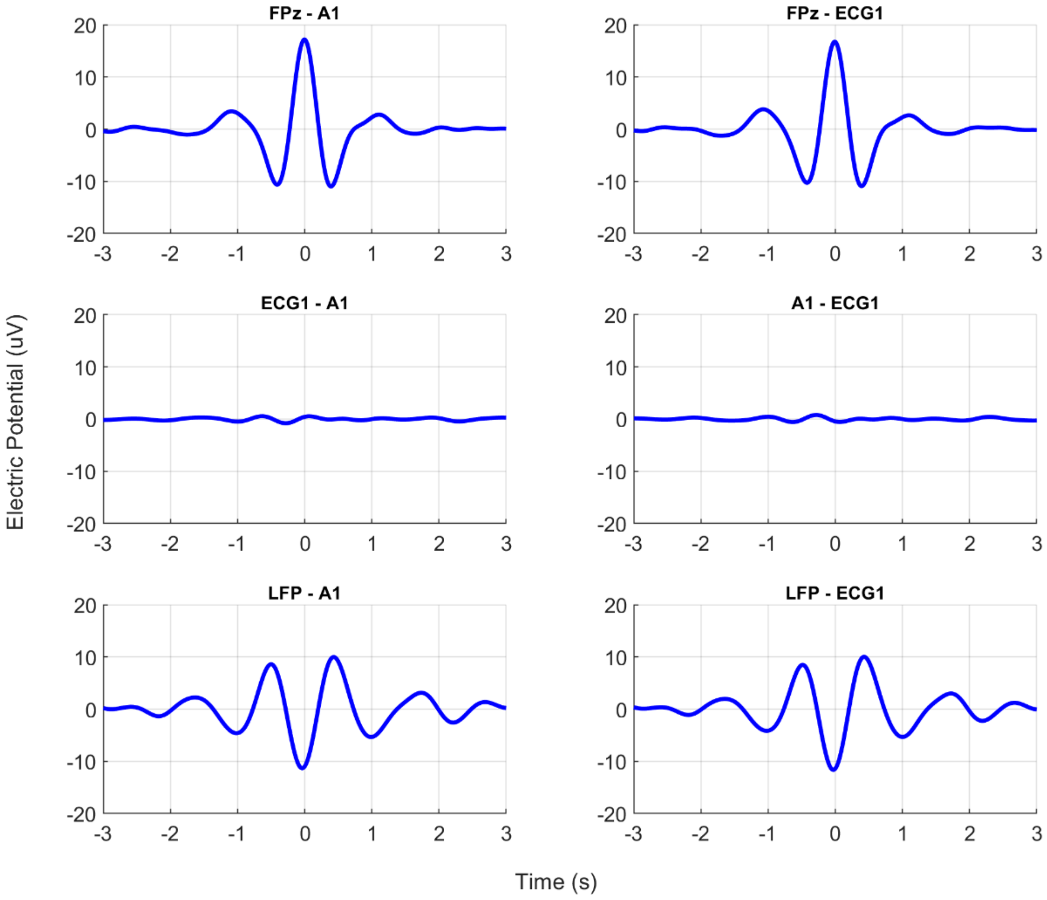


**Supplementary Fig SWMC. Mastoid electrode does not capture activity time-locked to frontal slow waves**. Example of time-locked slow waves (peak detected offline in the Fpz referenced to joined mastoid, time=0 in subplots, patient 1, Recoding Session 1) for FPz and STN-LFP signals referenced to A1 (left mastoid, left column) and the ECG1 (cardiac electrode located on chest, right column).

#### 3.2. Event-related potential analysis

We performed exploratory ERP and time-frequency analysis of response to the auditory stimuli presentation in stim ON windows and compared it to the time-locked waves obtained for other types of PTAS windows. As shown in Supplementary Fig. ERP1 below, when comparing the Fpz ERPs for the stim ON windows with spontaneous slow waves in sham ON windows, there was a significant change in ERP amplitude due to the PTAS (Fpz, mastoid reference, left column). Fpz ERP waveform is resembling a classical K-complex (Fpz, mastoid reference, right column). This finding is in line with the previously published works on PTAS response during sleep (Ngo et al., 2013; Krugliakova et al., 2020). The difference ERP waveform had pronounced slow large-amplitude deflection at the latencies of approximately 700 ms and 1200 ms, which might be considered a classic N550 and P900 K-complex components (Laurino et al., 2014) delayed due to the latency shift observed in older individuals (Bennett et al., 2004). For the mastoid-referenced STN-LFP, we also observed a high-amplitude time-locked response to PTAS (STN-LFP, mastoid reference, left column). Of note, as compared to the surface EEG, N550 and P900 components in the STN-LFP were on a similar latency, but the polarity of the waveform was reversed, confirming previous findings in intracranial recordings (Botella-Soler et al., 2012). A detectable ERP could be observed also for the two other reference montages for STN-LFP (bipolar and Laplacian).

The prestimulus interval (-0.5 to 0 s) could also be of interest for the purpose of our study, as a positive or negative deflection found in the prestimulus interval indicates that the phase of the slow wave was consistent across averaged trials. Thus, as expected from the performance of the PTAS algorithm, there was a negative deflection in the prestimulus for all types of windows in the mastoid-referenced Fpz signal. For the mastoid-referenced STN-LFP, there was also a consistent response, but the polarity reversed. In the case of bipolar STN-LFP, there was no distinguishable wave in the prestimulus. Finally, for the Laplacian-referenced STN-LFP, there was a slight negative deflection, indicating that the phase of the surface EEG potentially could be detected with this type of reference montage for STN-LFP. The latter observation confirms the finding presented in Fig. 6, showing significant coherence between Fpz and Laplacian-referenced STN-LFP.

Time-frequency analysis (Supplementary Fig. ERP1, right column) of the Fpz data demonstrates a well-described pattern of theta-sigma (5-6 Hz and 9-14 Hz) response following stimulus presentation (Lehmann et al., 2016; Leminen et al., 2017; Papalambros et al., 2019). A similar response was present in these frequency bands for all three types of STN-LFP referencing, with the most alike pattern for Laplacian-referenced STN-LFP. This result agrees with the power changes presented on the spectrogram in Fig. 4.

Next, we performed the ERP analysis of the Fpz and bipolar-referenced STN-LFP for the DBS+ and DBS- Sessions (Supplementary Fig. ERP23). The Fpz ERP was similar for DBS+ and DBS- Sessions and had N550 and P900 components. The STN-LFP signal also revealed a significant amplitude change at the latency of N550 and P900 components, which was more pronounced in DBS- Session. The time-frequency analysis of the DBS+ and DBS- Sessions has shown an auditory-stimulation evoked increase in theta and sigma frequency ranges.

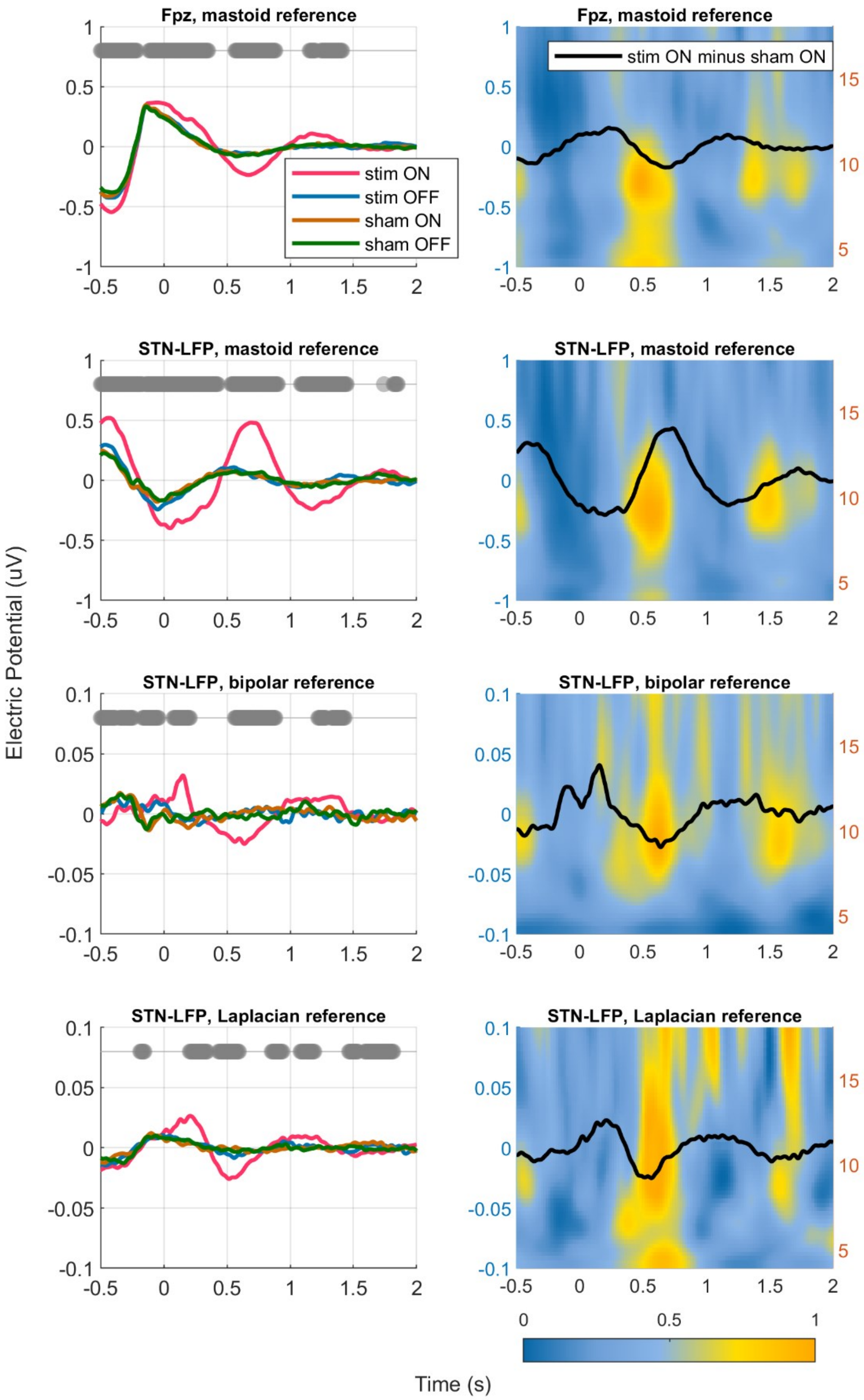


**Supplementary Fig ERP1. Event-related potentials and time-frequency representations for the Recording Session 1.** Data is presented for the mastoid-referenced Fpz (N=5) and STN-LFP referenced in three different ways (mastoid, bipolar, and Laplacian reference montages, N=10, mean±CI). Zero on the time axis corresponds to the moment of auditory stimulus presentation (or trigger in stim OFF and both sham windows). The left column shows ERPs for four types of windows for PTAS (color-coded). Gray dots indicate p<0.05 for stim ON vs. sham ON comparison, not cluster corrected. The right column row shows power changes following the stimulus presentation (stim ON/sham ON) overlayed with a difference waveform (stim ON minus sham ON).

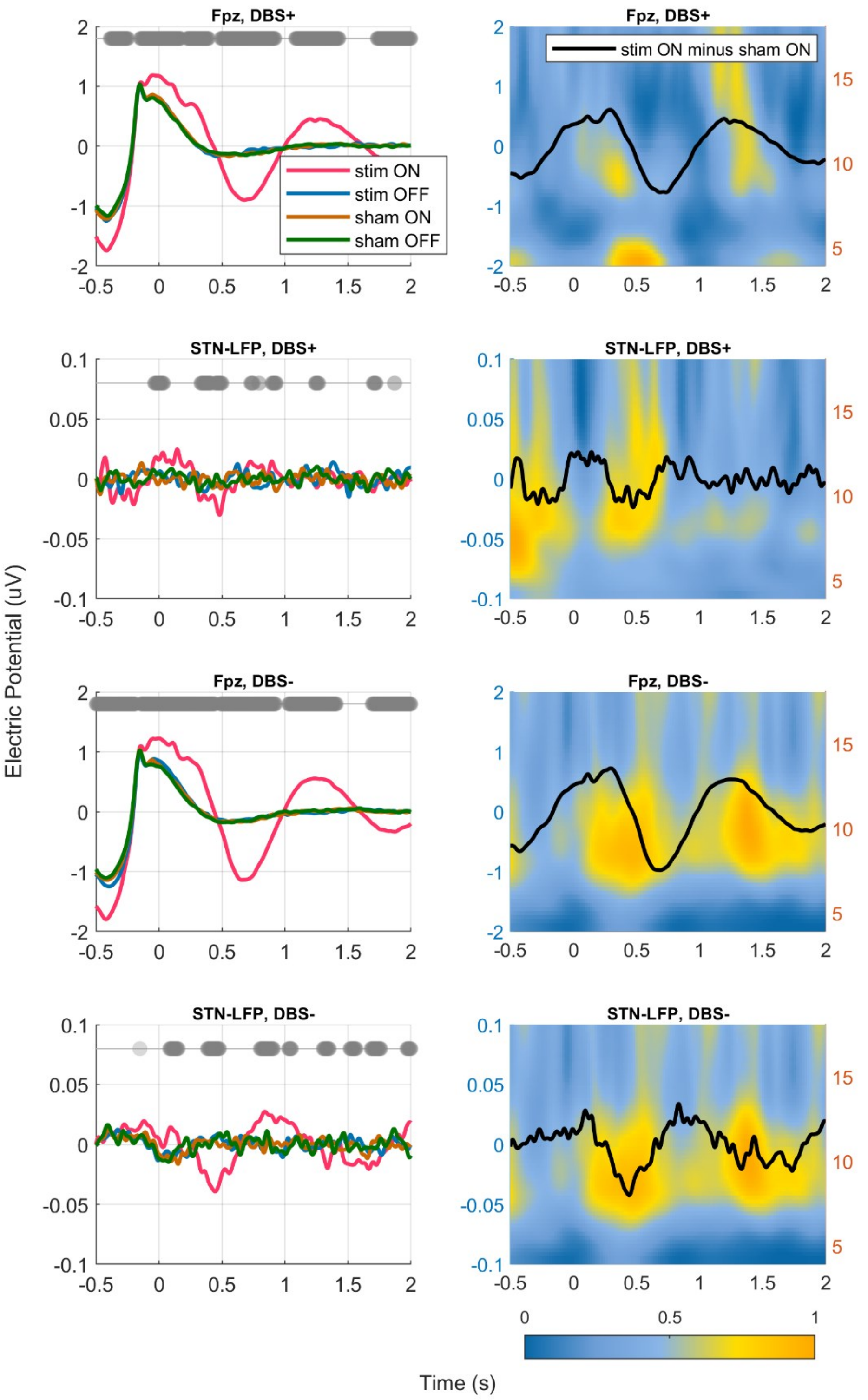


**Supplementary Fig ERP23. Event-related potentials and time-frequency representations DBS+ and DBS- Recording Sessions.** Data is presented for the mastoid-referenced Fpz (N=5) and bipolar-referenced STN-LFP (N=10, mean±CI). Zero on the time axis corresponds to the moment of auditory stimulus presentation (or trigger in stim OFF and both sham windows). The left column shows ERPs for four types of windows for PTAS (color-coded). Gray dots indicate p<0.05 for stim ON vs. sham ON comparison, not cluster corrected. The right column row shows power changes following the stimulus presentation (stim ON/sham ON) overlayed with a difference waveform (stim ON minus sham ON). Of note, there was a hardware highpass filter at 1 Hz applied to the Percept PC neurostimulator recording.

### 4. Supplementary references


Bennett, I. J., Golob, E. J., and Starr, A. (2004). Age-related differences in auditory event-related potentials during a cued attention task. *Clin. Neurophysiol.* 115, 2602–2615. doi:10.1016/j.clinph.2004.06.011.

Berry, R., Brooks, R., Gamaldo, C., Harding, S., Lloyd, R., Marcus, C., et al. (2015). *The AASM Manual for the Scoring of Sleep and Associated Events*. Darien, Illinois: American Academy of Sleep Medicine.

Botella-Soler, V., Valderrama, M., Crépon, B., Navarro, V., and van Quyen, M. Le (2012). Large-scale cortical dynamics of sleep slow waves. *PLoS One* 7, 1–10. doi:10.1371/journal.pone.0030757.

Ferster, M. L., Da Poian, G., Menachery, K., Schreiner, S. J., Lustenberger, C., Maric, A., et al. (2022). Benchmarking Real-Time Algorithms for In-Phase Auditory Stimulation of Low Amplitude Slow Waves With Wearable EEG Devices During Sleep. *IEEE Trans. Biomed. Eng.* 69, 2916–2925. doi:10.1109/TBME.2022.3157468.

Krugliakova, E., Volk, C., Jaramillo, V., Sousouri, G., and Huber, R. (2020). Changes in cross-frequency coupling following closed-loop auditory stimulation in non-rapid eye movement sleep. *Sci. Rep.* 10. doi:10.1038/s41598-020-67392-w.

Laurino, M., Menicucci, D., Piarulli, A., Mastorci, F., Bedini, R., Allegrini, P., et al. (2014). Disentangling different functional roles of evoked K-complex components: Mapping the sleeping brain while quenching sensory processing. *Neuroimage* 86, 433–445. doi:10.1016/j.neuroimage.2013.10.030.

Lehmann, M., Schreiner, T., Seifritz, E., and Rasch, B. (2016). Emotional arousal modulates oscillatory correlates of targeted memory reactivation during NREM, but not REM sleep. *Sci. Rep.* 6, 39229. doi:10.1038/srep39229.

Leminen, M. M., Virkkala, J., Saure, E., Paajanen, T., Zee, P. C., Santostasi, G., et al. (2017). Enhanced memory consolidation via automatic sound stimulation during non-REM sleep. *Sleep* 40. doi:10.1093/sleep/zsx003.

Lustenberger, C., Ferster, M. L., Huwiler, S., Brogli, L., Werth, E., Huber, R., et al. (2022). Auditory deep sleep stimulation in older adults at home: a randomized crossover trial. *Commun. Med.* 2. doi:10.1038/s43856-022-00096-6.

Ngo, H. V. V., Martinetz, T., Born, J., and Mölle, M. (2013). Auditory closed-loop stimulation of the sleep slow oscillation enhances memory. *Neuron* 78, 545–553. doi:10.1016/j.neuron.2013.03.006.

Papalambros, N. A., Weintraub, S., Chen, T., Grimaldi, D., Santostasi, G., Paller, K. A., et al. (2019). Acoustic enhancement of sleep slow oscillations in mild cognitive impairment. *Ann. Clin. Transl. Neurol.* doi:10.1002/acn3.796.
